# Monomeric CRP and Negative Acute Phase Proteins but not pentameric CRP are biomarkers of major depression and especially major dysmood disorder

**DOI:** 10.1101/2025.09.03.25334980

**Authors:** Abbas F. Almulla, Mengqi Niu, Drozdstoy Stoyanov, Yingqian Zhang, Michael Maes

## Abstract

**Background:** Contrary to the negative acute-phase protein (APP) response, there is no consistent correlation between serum pentameric C-reactive protein (pCRP) and major depression (MDD). Monomeric CRP (mCRP), a dissociation product of pCRP under immune-inflammatory conditions, exhibits pro-inflammatory effects; however, it has not been investigated in MDD or its subtypes, major dysmood disorder (MDMD) and simple dysmood disorder (SDMD).

**Objective:** To examine serum mCRP, albumin, transferrin, M1 macrophage and Thelper-17 immune profiles, and adverse childhood experiences (ACEs) in MDD, MDMD and SDMD.

**Methods:** Seventy-nine MDMD patients, 30 SDMD patients, and 40 controls were included. Serum mCRP was measured by ELISA; albumin, transferrin, and pCRP by biochemical assays; and cytokines using Luminex technology.

**Results:** MDMD patients showed significantly higher mCRP compared with SDMD and controls, while both patient groups exhibited reduced albumin and transferrin. Combining mCRP with albumin and transferrin showed an adequate accuracy for MDD (area under the ROC Curve = 0.793). Adding IL-17A and ACEs improved accuracy (ROC=0.855). Serum mCRP levels are additionally associated with pCRP, M1 macrophage profile, body mass index, and ACEs. Up to 36.6% of the variance in overall severity of depression was explained by mCRP, T-helper-17 profile, ACEs (all positively), albumin and transferrin (both inversely).

**Conclusion:** Future research in MDD should employ mCRP rather than pCRP as a biomarker of depression/MDMD. Combining mCRP with biomarkers of the negative AP response shows that around 63.7% of MDD patients have a smoldering acute phase response with high specificity.

## Introduction

Major depressive disorder (MDD) is increasingly identified with disruptions in neuroimmune, metabolic, and oxidative (NIMETOX) pathways (Maes, Almulla et al. 2025). MDD in its various stages is linked to the upregulated immune-inflammatory response system (IRS), particularly the M1 macrophage profile and T helper (Th)-1 and Th-17 pathways, as evidenced by increased levels of interleukin (IL)-1β, IL-6, IL-17A, and tumor necrosis factor (TNF)-α (Maes, Almulla et al. 2025). The compensatory immunoregulatory system (CIRS), which includes anti-inflammatory cytokines such as IL-10 and IL-4, as well as sIL-1RA, is frequently reduced (Maes and Carvalho 2018, Maes, Almulla et al. 2025). The disparity between IRS and CIRS contributes to chronic inflammation and neurotoxicity, which are key characteristics of the acute phase of MDD (Maes, Almulla et al. 2025).

A precision nomothetic psychiatry approach has refined this model by classifying MDD into two primary subtypes, namely major dysmood disorder (MDMD) and simple dysmood disorder (SDMD) (Maes, Rachayon et al. 2022). Patients with MDMD exhibit greater severity of depression, anxiety, chronic fatigue, and suicidal behaviors, along with increased recurrence of illness (ROI) indices, in comparison to those with SDMD (Maes, Rachayon et al. 2022, Almulla, Abbas Abo Algon et al. 2024). MDMD is biologically defined by significant IRS and NIMETOX activation, which includes increased M1 macrophage and Th-1/Th-17 signaling (Maes, Almulla et al. 2025). In contrast, the initial episode of SDMD is more closely linked to CIRS inhibition (Maes, Rachayon et al. 2022, Maes, Vasupanrajit et al. 2023, Maes, Vasupanrajit et al. 2025). Immune activation in first-episode MDMD is significantly associated with adverse childhood experiences (ACEs), highlighting the impact of early-life stress on immune-inflammatory dysregulation (Almulla, Algon et al. 2024).

The acute phase response is part of IRS activation. In this process, hepatocytes elevate the levels of positive acute phase proteins (APPs), including C-reactive protein (CRP), while the levels of negative APPs, particularly albumin and transferrin, decrease (Maes 1993, Almulla, Thipakorn et al. 2022, Maes, Niu et al. 2025). Reduced levels of albumin and transferrin are consistently observed in MDD and exhibit a strong correlation with immune activation, ACEs, and the severity of physio-affective symptoms (Maes 1993, Maes, Niu et al. 2025). Consequently, these markers offer a more direct measure of the acute phase response in MDD as compared with measurements of serum pentameric CRP (Maes, Niu et al., 2025).

Nevertheless, pentameric CRP (pCRP) is frequently suggested as a biomarker for “inflammatory depression” when serum levels surpass 3 or 5 mg/L (Raison, Rutherford et al. 2013, Wessa, Janssens et al. 2024, Jha, Leboyer et al. 2025). Certain authors have proposed utilizing pCRP thresholds to inform treatment decisions regarding anti-inflammatory agents (Raison, Rutherford et al. 2013, Wessa, Janssens et al. 2024). Nevertheless, the elevation of pCRP is more closely linked to obesity, metabolic syndrome (MetS), and ACEs than to MDD itself (Douglas, Taylor et al. 2004, Moraes, Maes et al. 2017, Khan, Leonard et al. 2020, Almulla, Kitov et al. 2025, Maes, Niu et al. 2025). Moraes et al. (2017) established that a large part (50%) of the variance in pCRP is determined by age, BMI and ACEs, namely sexual abuse (Moraes, Maes et al. 2017). Recently, our research indicated that in drug-naïve obese patients with MetS, depressive symptoms are not associated with pCRP but are instead related to other metabolic and immune indices (Almulla, Kitov et al. 2025). In addition, Maes et al. revealed no association between MDD and pCRP, whilst lowered albumin and transferrin significantly explained MDD (Maes, Niu et al. 2025). These findings confirm the role of negative APPs as biomarkers of MDD and pCRP as a biomarker of metabolic conditions (Almulla, Kitov et al. 2025, Maes, Niu et al. 2025).

CRP exists in various conformations, pCRP and monomeric CRP (mCRP). The latter is produced via the conformational dissociation of pCRP under certain conditions at sites of inflammation and tissue injury, where it demonstrates significant pro-inflammatory effects (Thiele, Habersberger et al. 2014, Olson, Hornick et al. 2023). mCRP activates endothelial cells, leukocytes, and platelets, induces oxidative stress, and promotes the release of inflammatory mediators, including IL-1, IL-6, IL-8, and TNF-α (Molins, Peña et al. 2011, Ruiz-Fernández, Gonzalez-Rodríguez et al. 2021, Olson, Hornick et al. 2023). Elevated mCRP has been associated with cardiovascular disease, Alzheimer’s disease, systemic lupus erythematosus, and hepatitis, showing stronger correlations with disease severity compared to pCRP (Jakuszko, Krajewska et al. 2017, Caprio, Badimon et al. 2018, Hornick and Potempa 2023, Gao, Yuan et al. 2024). Due to its proinflammatory effects, mCRP has been suggested as a potential biomarker for MDD (Hornick and Potempa 2023), however, no previous research has assessed mCRP.

This study is the first to examine serum mCRP levels in combination with pCRP, negative APPs (albumin and transferrin), M1 macrophage, Th-17 and CIRS profiles in MDMD and SDMD versus healthy controls. We will also investigate whether the integration of mCRP with negative APPs and inflammatory profiles yields enhanced diagnostic and explanatory value for distinct subtypes of MDD and severity of illness. Our hypothesis is that mCRP is significantly increased in MDD and its combination with albumin and transferrin will significantly enhance its ability to predict MDD rather than pCRP.

## Methods

### Subjects

This study included 165 participants consisting of 79 patients with MDMD, 30 with SDMD and 40 healthy controls. This case–control cross-sectional study was performed at the Psychiatric Center of Sichuan Provincial People’s Hospital, Chengdu, China. Participants were between 18 and 65 years of age, and both sexes were represented. MDD patients were diagnosed according to DSM-5, and eligibility required a Hamilton Depression Rating Scale (HAMD-21) score greater than 18 (Hamilton 1960). Furthermore, patients were classified into MDMD (the most severe subtype) and SDMS (milder depression) based on established criteria from prior research (Maes, Rachayon et al. 2022). Controls were recruited from hospital staff, their relatives, and acquaintances of patients, and they were matched to cases by age, sex, education, and body mass index (BMI). Written informed consent was obtained from all participants or their legal guardians. Ethical approval was granted by the Research Ethics Committee of the Sichuan Provincial People’s Hospital [Ethics (Research) 2024-203].

In the present study, the exclusion criteria includes: a) other psychiatric disorders rather than MDD such as schizophrenia, schizoaffective disorder, bipolar disorder, psycho-organic disorders, autism spectrum disorders, substance use disorders apart from nicotine dependence; b) systemic medical diseases including autoimmune disorders, systemic lupus erythematosus, inflammatory bowel disease, psoriasis, type 1 diabetes mellitus, rheumatoid arthritis, chronic obstructive pulmonary disease, or cancer; c) severe allergic reaction during the past month; d) pregnancy or breastfeeding; e) developmental or personality disorders, or neurological illnesses such as stroke, epilepsy, brain tumors, Parkinson’s disease, Alzheimer’s disease, or multiple sclerosis; f) infection within the last three months; g) current use of immunosuppressants, glucocorticoids, or other immunomodulators; h) therapeutic doses of antioxidants or omega-3 supplements within the last three months; i) recent surgery within the last three months; or j) frequent use of analgesics. In addition, we excluded controls if they had a current or past diagnosis of MDD, dysthymia, DSM-IV anxiety disorders, or a family history of affective disorders, suicide, or substance use disorders (other than nicotine dependence).

### Clinical Assessments

A trained physician conducted semi-structured interviews that recorded demographic data, illness course, medical and psychiatric history, and family history. Psychiatric diagnoses were confirmed using the Mini International Neuropsychiatric Interview (M.I.N.I.) (Sheehan, Lecrubier et al. 1998), which assessed depressive episodes, dysthymia, hypomanic episodes, suicidal ideation, panic disorder, agoraphobia, social anxiety disorder, generalized anxiety disorder, obsessive–compulsive disorder, post-traumatic stress disorder, alcohol and non-alcoholic substance dependence/abuse, psychotic disorders, eating disorders, and antisocial personality disorder.

Symptom severity was assessed on the same day with the 21-item Hamilton Depression Rating Scale (HAMD) (Hamilton 1960), the Hamilton Anxiety Rating Scale (HAMA) (Hamilton 1959), and the State version of the State–Trait Anxiety Inventory (STAI) (Spielberger, Gorsuch et al. 1983). A principal component (PC) was extracted from the HAMD, HAMA, and STAI values which explained 83.61% of the variance. This PC analysis showed a Kaiser-Meyer-Olkin metric of 0.727 (p<0.0001), and all three scores were highly loaded on the first PC (>0.85) with a Cronbach’s Alpha= 0.902. This PC score was employed as an overall severity of depression (OSOD) index. ACEs were assessed using the Childhood Trauma Questionnaire–Short Form (CTQ-SF) (Bernstein, Stein et al. 2003), validated in Chinese by Zhao Xingfu (Zhao 2005). Emotional and physical abuse, sexual abuse, emotional neglect, and physical neglect scores were calculated following prior work (Vasupanrajit, Maes et al. 2024). Since sexual abuse appears to be a specific significant predictor of pCRP (Moraes, Maes et al. 2017) we used sexual abuse and the sum of the four other ACEs in the analyses (labeled as “sum four ACEs”).

### Anthropometrics and Metabolic Syndrome

Measurements included weight, height, waist circumference (WC), and BMI. The calculation of BMI involves dividing weight in kilograms by the square of height in meters. WC was taken midway between the iliac crest and the lowest rib. A composite adiposity index was computed as zBMI + zWC. Metabolic syndrome (MetS) was defined according to the 2009 Joint Scientific Statement of the American Heart Association and National Heart, Lung, and Blood Institute (Alberti, Eckel et al. 2009), requiring at least three of the following: WC ≥ 90 cm in men or ≥ 80 cm in women, triglycerides ≥ 150 mg/dL, HDL-cholesterol < 40 mg/dL in men or < 50 mg/dL in women, systolic blood pressure ≥ 130 mm Hg or diastolic ≥ 85 mm Hg or antihypertensive treatment, and fasting glucose ≥ 100 mg/dL or a diagnosis of diabetes. MetS ranking was based on the number of fulfilled criteria.

### Blood Collection and Assays

Between 06:30 and 08:00 a.m., fasting venous blood (30 mL) was drawn into serum tubes with disposable syringes. Samples underwent centrifugation at 3500 rpm, after which serum was aliquoted into Eppendorf tubes and preserved at −80 °C until analysis. mCRP concentrations were measured with a BioVendor ELISA kit (Brno, Czech Republic), sensitivity 0.63 ng/mL, intra-assay CV < 10%, and inter-assay CV < 15%. pCRP was quantified using a particle-enhanced immunoturbidimetric assay (DIAYS DIAGNOSTIC SYSTEM, Shanghai) on an ADVIA 2400 analyzer (Siemens Healthcare Diagnostics Inc). The assay had sensitivity 0.3 mg/L, intra-assay CV 2.80%, and inter-assay CV 2.17%. Serum transferrin was determined by immunoturbidimetry (DIAYS DIAGNOSTIC SYSTEM) on the ADVIA 2400, sensitivity 0.03 g/L, intra-assay CV 1.96%, and inter-assay CV 0.67%. Albumin was measured with a Bromocresol Green kit (Beijing Strong Biotechnologies, Inc.) on the ADVIA 2400, intra-assay CV 1.20% and inter-assay CV 2.10%. A negative acute-phase index was calculated as z albumin + z transferrin.

Cytokines were analyzed using Luminex xMAP technology on the Luminex 200 system (Luminex Corporation, Austin, TX, USA) with the Human XL Cytokine Fixed Panel (bio-techne, R&D Systems; Cat. No. LKTM014B). Fluorescence intensity (FI) and concentrations were obtained for 46 cytokines and chemokines with FI values corrected for blanks. The protocol included: (1) dilute samples two-fold with Calibrator Diluent RD6-65; (2) add 50 μL diluted sample and 50 μL microparticle cocktail per well, shake 850 rpm for 2 h at room temperature; (3) wash three times, add 50 μL Biotin-Antibody Cocktail, incubate 1 h at 850 rpm; (4) wash again, add 50 μL Streptavidin-PE, incubate 30 min at 850 rpm; (5) final wash and resuspension in 100 μL buffer for 2 min per well. The Luminex 200 quantified all analytes. Intra-assay CVs were < 5% and inter-assay CVs < 11.2%. Indices were created for immune activity: M1 macrophage activity was the z-score sum of z IL-1 + z IL-6 + z TNF-α + z sIL-1RA (Maes and Carvalho 2018); CIRS activity was the z-score sum of z IL-4 + z IL-10 + z EGF (Maes and Carvalho 2018) ; and Th-17 activity was defined as z IL-6 + z IL-17A (Maes and Carvalho 2018).

### Data Analysis

This research employed IBM SPSS for Windows, version 30, to perform all statistical analysis. Contingency tables tested categorical associations, while analysis of variance (ANOVA) compared continuous outcomes. Multiple comparisons were adjusted with the false discovery rate (FDR). Pearson correlations examined associations among continuous variables, and point-biserial correlations between continuous and binary variables. Binary logistic regression models contrasted MDD with controls, and MDMD with SDMD. Diagnosis (MDD or MDMD) served as the dependent variable, with controls or SDMD as reference. Covariates included age, sex, BMI, and smoking status. Results included Nagelkerke pseudo-R², Wald statistics with p-values, odds ratios with 95% CIs, and regression coefficients (B) with standard errors (SE). The Wald statistic was defined as (B/SE)². To address subgroup imbalance (SDMD and controls are underrepresented compared with MDD and MDMD, respectively), oversampling (random replicate) was applied for SDMD versus MDMD and for controls versus MDD. Cross-validated classification accuracy was tested with linear discriminant analysis and 10-fold cross-validation. Model performance was summarized with the area under the ROC (receiver operating characteristic) curve (AUC), Gini index, maximum Kolmogorov–Smirnov (Max K–S) values, and overall fit. Multiple linear regression analyses predicted acute-phase proteins or rating scale scores from demographic variables, metabolic indicators, and biomarkers including ACEs. Both manual and automated stepwise procedures were employed. Automated modeling applied entry and removal thresholds of p = 0.05 and p = 0.07, respectively. Results included standardized beta coefficients, degrees of freedom, p-values, R², and F-statistics. Heteroskedasticity was tested with the White and modified Breusch–Pagan tests. Multicollinearity was checked by tolerance and variance inflation factors. All analyses were two-tailed with α = 0.05, and data transformations (log10, square-root, rank-order, or Winsorizing) were applied where required.

## Results

### Socio-demographic, clinical and biomarkers data

Sociodemographic and clinical characteristics for MDMD, SDMD, and controls are presented in **Table 1**. There were no significant differences across groups in age, sex ratio, or smoking. Metabolic variables also did not differ significantly, including MetS prevalence, MetS ranking, waist circumference, and BMI. Total HAMD, HAMA, and STAI scores were higher in MDMD than SDMD, and both patient groups scored higher than controls. Transferrin and albumin were reduced in MDMD and SDMD compared with controls, with no differences between the two patient groups. M1 macrophages were higher in MDMD than SDMD; neither patient group differed from controls. T helper-17 was elevated in MDMD compared with SDMD and controls, with no difference between SDMD and controls. The CIRS profile was increased in MDMD relative to SDMD and controls, with no difference between SDMD and controls.

**Table 1.**
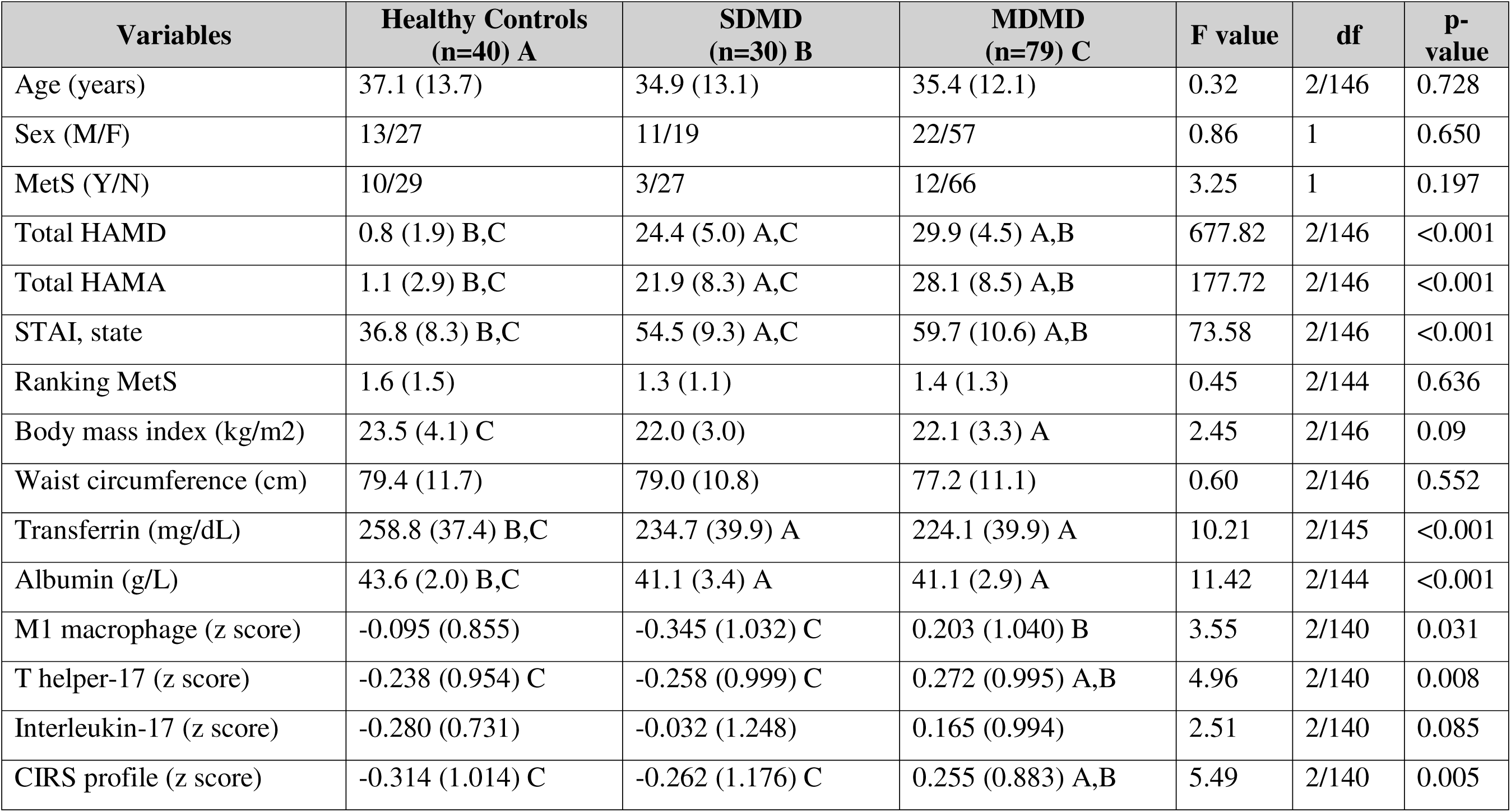
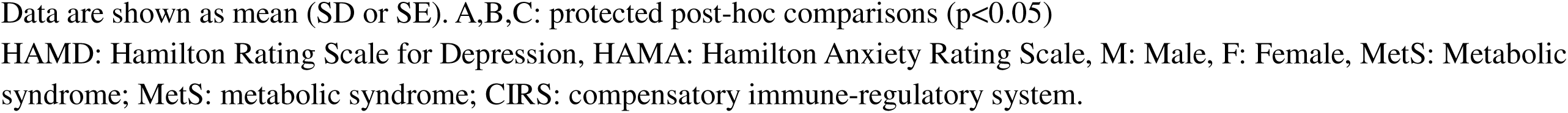
Socio-demographic, clinical and biomarker data of patients with major dysmood disorder (MDMD), simple dysmood disorder (SDMD) and healthy controls (HC).

| Variables | Healthy Controls<br>(n=40) A | SDMD<br>(n=30) B | MDMD<br>(n=79) C | F value | df | P-value |
| --- | --- | --- | --- | --- | --- | --- |
| Age (years) | 37.1 (13.7) | 34.9 (13.1) | 35.4 (12.1) | 0.32 | 2/146 | 0.728 |
| Sex (M/F) | 13/27 | 11/19 | 22/57 | 0.86 | 1 | 0.650 |
| MetS (Y/N) | 10/29 | 3/27 | 12/66 | 3.25 | 1 | 0.197 |
| Total HAMD | 0.8 (1.9) B,C | 24.4 (5.0) A,C | 29.9 (4.5) A,B | 677.82 | 2/146 | <0.001 |
| Total HAMA | 1.1 (2.9) B,C | 21.9 (8.3) A,C | 28.1 (8.5) A,B | 177.72 | 2/146 | <0.001 |
| STAI, state | 36.8 (8.3) B,C | 54.5 (9.3) A,C | 59.7 (10.6) A,B | 73.58 | 2/146 | <0.001 |
| Ranking MetS | 1.6 (1.5) | 1.3 (1.1) | 1.4 (1.3) | 0.45 | 2/144 | 0.636 |
| Body mass index (kg/m<sup>2</sup>) | 23.5 (4.1) C | 22.0 (3.0) | 22.1 (3.3) A | 2.45 | 2/146 | 0.09 |
| Waist circumference (cm) | 79.4 (11.7) | 79.0 (10.8) | 77.2 (11.1) | 0.60 | 2/146 | 0.552 |
| Transferrin (mg/dL) | 258.8 (37.4) B,C | 234.7 (39.9) A | 224.1 (39.9) A | 10.21 | 2/145 | <0.001 |
| Albumin (g/L) | 43.6 (2.0) B,C | 41.1 (3.4) A | 41.1 (2.9) A | 11.42 | 2/144 | <0.001 |
| M1 macrophage (z score) | -0.095 (0.855) | -0.345 (1.032) C | 0.203 (1.040) B | 3.55 | 2/140 | 0.031 |
| T helper-17 (z score) | -0.238 (0.954) C | -0.258 (0.999) C | 0.272 (0.995) A,B | 4.96 | 2/140 | 0.008 |
| Interleukin-17 (z score) | -0.280 (0.731) | -0.032 (1.248) | 0.165 (0.994) | 2.51 | 2/140 | 0.085 |
| CIRS profile (z score) | -0.314 (1.014) C | -0.262 (1.176) C | 0.255 (0.883) A,B | 5.49 | 2/140 | 0.005 |

Data are shown as mean (SD or SE). A,B,C: protected post-hoc comparisons ( $p < 0.05$ )
HAMD: Hamilton Rating Scale for Depression, HAMA: Hamilton Anxiety Rating Scale, M: Male, F: Female, MetS: Metabolic syndrome; MetS: metabolic syndrome; CIRS: compensatory immune-regulatory system.

**Table 2** shows no significant differences in pCRP across groups. In contrast, mCRP was higher in MDMD than controls, while SDMD and controls did not differ. In two other models, mCRP remained higher in MDMD than SDMD or controls after covarying for different explanatory variables, including pCRP, which was a significant predictor of mCRP (F = 12.57, df = 2/142, p < 0.001).

**Table 2.** Measurements of pentameric and monomeric C-reactive protein (CRP) in patients with major dysmood disorder (MDMD), simple dysmood disorder (SDMD) and healthy volunteers (HC)

| Variables | HC<br>(n=40) A | SDMD<br>(n=30) B | MDMD<br>(n=79) C | F value | df | p-value |
| --- | --- | --- | --- | --- | --- | --- |
| Pentameric CRP*^ | 0.992 (1.120) | 2.488 (1.072) | 2.212 (0.657) | 0.288 | 2/142 | 0.592 |
| Momomeric CRP*^ | 1.091 (0.069) C | 1.284 (0.066) | 1.410 (0.040)A | 9.92 | 2/142 | 0.002 |
| Residualized monomeric Log CRP*^ | 0.044 (0.017) C | 0.094 (0.018) | 0.129 (0.011) A | 8.14 | 2/142 | <0.001 |
| Residualized monomeric Log CRP** | 0.050 (0.017) C | 0.091 (0.017) | 0.128 (0.011) A | 7.50 | 2/142 | <0.001 |

All data are shown are marginal estimated mean (SE). A,B,C: protected post-hoc comparisons ( $p < 0.05$ ). \* Processed in Log10 transformation; ^: adjusted for body mass index and drug status (age and sex are not significant); \*\* After adjusting for body mass index, medication status, and pentameric CRP (the latter was highly significant as covariate, namely: $F=12.57$ , $df=2/142$ , $p < 0.001$ ).

### Correlations between pCRP, mCRP, metabolic indices, and ACEs

**Table 3** presents Pearson’s product–moment correlations between pCRP and mCRP values and clinical measures (MetS, MetS rank, BMI, waist circumference) and psychological stressors (sexual abuse and the sum four ACEs). pCRP showed significant positive correlations with the metabolic variables and sexual abuse, but not the sum four ACEs. mCRP was significantly and positively correlated with all variables except sexual abuse. In contrast, residualized mCRP was positively correlated with the sum four ACEs.

**Table 3.** Intercorrelation matrix among pentameric and monomeric C-reactive protein (CRP), metabolic variables and adverse childhood experiences (ACEs)

| Variables | Pentameric CRP | monomeric CRP | Residualized monomeric CRP<sup>^</sup> |
| --- | --- | --- | --- |
| Metabolic syndrome (MetS) | 0.260<sup>**</sup> (<0.001) | 0.164<sup>*</sup> (0.037) | 0.041 (0.601) |
| Rank MetS | 0.285<sup>**</sup> (<0.001) | 0.216<sup>**</sup> (0.006) | 0.059 (0.455) |
| Body mass index | 0.380<sup>**</sup> (<0.001) | 0.249<sup>**</sup> (0.001) | 0 (1) |
| Waist circumference | 0.345<sup>**</sup> (<0.001) | 0.208<sup>**</sup> (0.008) | 0.002 (0.979) |
| Sexual abuse | 0.161<sup>*</sup> (0.039) | 0.097 (0.219) | 0.140 (0.074) |
| Sum four ACEs | 0.143 (0.067) | 0.194<sup>*</sup> (0.013) | 0.253<sup>**</sup> (0.001) |

<sup>^</sup>After adjusting for body mass index, medication status, $p < 0.05$ , $p < 0.01$ .

### Accuracy of pCRP, mCRP and negative APPs

**Table 4** presents binary logistic regression analyses with the MDD diagnosis (healthy control as reference group) as the dependent variable and positive acute phase proteins (mCRP, pCRP), negative acute phase proteins (transferrin, albumin), and inflammatory biomarkers (IL-17, M1 macrophage) as explanatory variables. In Model #1, MDD was not significantly associated with pCRP. In Model #2, mCRP was positively associated with MDD (χ² = 13.512, df = 1, p < 0.001; Nagelkerke = 0.072), with an overall accuracy of 60.7% (sensitivity = 62.4%, specificity = 59.0%). Model #3 shows that the residualized mCRP (adjusted for BMI and drug state) was positively associated with MDD (χ² = 31.538, df = 4, p < 0.001; Nagelkerke = 0.168), with an accuracy of 62.8% (sensitivity = 62.5%, specificity = 63.2%). As shown in **Table 5**, the cross-validated accuracy was 58% with an area under the ROC curve of 0.616.

**Table 4:** Results of binary logistic regression analyses with major depression (MDD) or major dysmood disorder (MDMD) as dependent variables and controls or simple dysmood disorder (SDMD) as reference groups.

| Dichotomies |  | Explanatory Variables | B | SE | Wald (df=1) | p | OR | 95% CI |
| --- | --- | --- | --- | --- | --- | --- | --- | --- |
| **MDD**<br>*versus healthy controls* | **Model #1** | Pentameric CRP | -0.237 | 0.139 | 2.91 | 0.088 | 0.789 | 0.601; 1.036 |
|  | **Model #2** | Monomeric CRP | 0.357 | 0.131 | 5.04 | 0.025 | 1.428 | 1.046; 1.950 |
|  | **Model #3** | Monomeric CRP* | 0.519 | 0.150 | 11.979 | <0.001 | 1.681 | 1.253; 2.256 |
|  | **Model #4** | Monomeric CRP* | 0.351 | 0.160 | 4.81 | 0.028 | 1.421 | 1.038; 1.945 |
|  |  | Transferrin | -0.532 | 0.167 | 10.16 | 0.001 | 0.588 | 0.424; 0.815 |
|  |  | Albumin | -0.857 | 0.159 | 20.56 | <0.001 | 0.424 | 0.293; 0.615 |
|  | **Model #5** | Monomeric CRP* | 0.330 | 0.167 | 3.93 | 0.047 | 1.391 | 1.004; 1.928 |
|  |  | Transferrin | -0.543 | 0.171 | 10.10 | .001 | 0.581 | 0.416; 0.812 |
|  |  | Albumin | -0.868 | 0.195 | 19.73 | <0.001 | 0.420 | 0.286; 0.616 |
|  |  | Interleukin-17 | 0.481 | 0.195 | 6.10 | 0.014 | 1.617 | 1.104; 2.368 |
| **MDMD** *versus*<br>**SDMD** | **Model #6** | Monomeric CRP* | 0.528 | 0.221 | 5.68 | 0.017 | 1.696 | 1.098; 2.617 |
|  | **Model #7** | Monomeric CRP* | 0.795 | 0.272 | 8.52 | 0.004 | 2.215 | 1.298; 3.777 |
|  |  | M1 macrophage | 0.473 | 0.194 | 5.97 | 0.015 | 1.605 | 1.098; 2.347 |
|  |  | Transferrin | -0.550 | 0.221 | 6.18 | 0.013 | 0.577 | 0.374; 0.890 |

\*Monomeric CRP: residualized values after adjusting for body mass index and drug status.

**Table 5.**
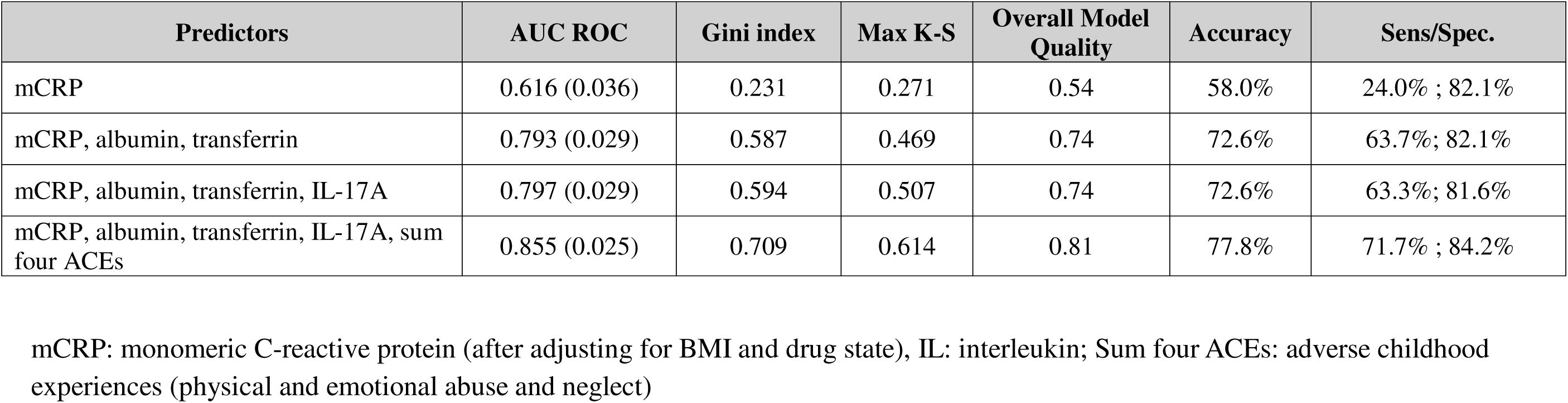
Diagnostic and predictive performance metrics of different models in differentiating major depressed patients from healthy controls.

| Predictors | AUC ROC | Gini index | Max K-S | Overall Model Quality | Accuracy | Sens/Spec. |
| --- | --- | --- | --- | --- | --- | --- |
| mCRP | 0.616 (0.036) | 0.231 | 0.271 | 0.54 | 58.0% | 24.0% ; 82.1% |
| mCRP, albumin, transferrin | 0.793 (0.029) | 0.587 | 0.469 | 0.74 | 72.6% | 63.7% ; 82.1% |
| mCRP, albumin, transferrin, IL-17A | 0.797 (0.029) | 0.594 | 0.507 | 0.74 | 72.6% | 63.3% ; 81.6% |
| mCRP, albumin, transferrin, IL-17A, sum four ACEs | 0.855 (0.025) | 0.709 | 0.614 | 0.81 | 77.8% | 71.7% ; 84.2% |

mCRP: monomeric C-reactive protein (after adjusting for BMI and drug state), IL: interleukin; Sum four ACEs: adverse childhood experiences (physical and emotional abuse and neglect)

In an additional regression (Model #4), MDD was associated with the residualized mCRP values (positive), transferrin and albumin (both negative) (χ² = 78.177, df = 6, p < 0.001; Nagelkerke = 0.379). Table 5 shows that the model accuracy was 76.1% (sensitivity = 75.8%, specificity = 76.3%), and the cross-validated accuracy was 72.6% (AUC = 0.793, Gini index = 0.587, Max K-S = 0.469). Analysis of the coordinates of the ROC curve showed that using these three APPs, 63.7% of the MDD patients were correctly classified with a specificity of 82.1%. Model #5 indicates that residualized mCRP and IL-17A (both positive), together with transferrin and albumin (both negative), are associated with MDD (χ² = 78.177, df = 6, p < 0.001; Nagelkerke = 0.062), with a cross-validated accuracy of 72.6% (AUC = 0.797, Gini index = 0.594, Max K-S = 0.507, see Table 5). Adding ACEs to the previous model resulted in a cross-validated accuracy of 77.8% (AUC = 0.855, Gini index = 0.709, Max K-S = 0.614).

**Table 3** also reports two models distinguishing MDMD (SDMD as reference group) using mCRP, M1 macrophage, and transferrin as predictors. Model #6 shows a positive association between mCRP and MDMD (χ² = 6.526, df = 1, p = 0.011; Nagelkerke = 0.062), with an accuracy of 55.4% (sensitivity = 36.7%, specificity = 80.0%). Model #7 shows a significant association of mCRP and M1 (both positive) and transferrin (negative) with MDMD (χ² = 24.968, df = 3, p = 0.011; Nagelkerke = 0.226), yielding an accuracy of 63.7% (sensitivity = 82.7%, specificity = 40.0%).

### Prediction of the severity of depression

**Table 6** summarizes the multivariate regression analyses in which pCRP, mCRP, and OSOD were used as dependent variables, with demographic data, negative APPs, ACEs, and inflammatory biomarkers entered as explanatory variables.

**Table 6.** Results of multiple regression analysis with serum C-reactive protein (CRP) and severity of depression score as dependent variables and biomarkers and clinical data as explanatory variables.

| Dependent Variables | Explanatory Variables | Coefficient statistics |  |  | Model statistics |  |  |  |
| --- | --- | --- | --- | --- | --- | --- | --- | --- |
| | | $\beta$ | t | p | R<sup>2</sup> | F | df | p |
| #1. Pentameric CRP | **Model** |  |  |  | 0.263 | 10.85 | 5/152 | <0.001 |
|  | Body mass index | 0.349 | 4.83 | <0.001 |  |  |  |  |
|  | Sexual abuse | 0.138 | 1.85 | 0.066 |  |  |  |  |
|  | M1 macrophage profile | 0.344 | 3.69 | <0.001 |  |  |  |  |
|  | CIRS profile | -0.241 | -2.62 | 0.010 |  |  |  |  |
|  | Sum four ACEs | 0.156 | 2.09 | 0.039 |  |  |  |  |
| #2. Monomeric CRP | **Model** |  |  |  | 0.160 | 9.78 | 3/154 | <0.001 |
|  | Body mass index | 0.268 | 3.55 | <0.001 |  |  |  |  |
|  | Sum four ACEs | 0.244 | 3.28 | 0.001 |  |  |  |  |
|  | M1 macrophage profile | 0.176 | 2.34 | 0.020 |  |  |  |  |
|  | Pentameric CRP | 0.219 | 2.89 | 0.004 |  |  |  |  |
| #3. Overall severity of depression | **Model** |  |  |  | 0.366 | 16.48 | 5/143 | <0.001 |
|  | Sum four ACEs | 0.376 | 5.32 | <.001 |  |  |  |  |
|  | Albumin | -0.177 | -2.40 | 0.018 |  |  |  |  |
|  | Monomeric CRP | 0.184 | 2.69 | 0.008 |  |  |  |  |
|  | Transferrin | -0.184 | -2.58 | 0.011 |  |  |  |  |
|  | T helper-17 | 0.136 | 2.02 | 0.046 |  |  |  |  |

CIRS: compensatory immune-regulatory system; Sum four ACEs: adverse childhood experiences (physical and emotional abuse and neglect)

Regression model #1 shows that 26.3% of the variance in pCRP was explained by BMI, sexual abuse, M1 macrophage, and four ACEs (all positive associations), together with CIRS (negative association). Regression model #2 demonstrates that 16% of the variance in mCRP was explained by BMI, four ACEs, M1 macrophage, and pCRP (all positive associations). Regression model #3 indicates that 36.6% of the variance in the OSOD score was explained by sum four ACEs, Th-17 profile, and mCRP (positive associations), along with albumin and transferrin (negative associations).

## Discussion

### Elevated mCRP in MDD

This study is the first to quantify serum mCRP in MDD. The first major finding of this study is that MDMD patients display significantly higher mCRP levels compared with SDMD and controls, while SDMD does not differ from controls. These differences remained significant after adjustments for BMI, medication status, age, and sex. In contrast, pCRP exhibited no differences between groups, corroborating previous studies that did not identify elevated pCRP (either raw or adjusted values) in MDD (Maes, Scharpe et al. 1992). The current study confirmed that, in addition to mCRP elevations, there were reductions in the negative APPs, specifically albumin and transferrin, among patients with MDD subtypes, thereby supporting the role of the negative APP response (Maes et al., 1991). This is consistent with earlier research indicating lower levels of albumin and transferrin in MDD (Ambrus and Westling 2019, Gregg, Carmody et al. 2020, Al-Marwani, Batieha et al. 2023) and meta-analytic data showing diminished albumin levels in MDD and bipolar disorder (Almulla, Thipakorn et al. 2022). The immune-inflammatory changes were most significant in MDMD, which exhibited greater mCRP levels, M1 macrophage activation, elevated Th-17 levels, and increased CIRS activity relative to SDMD. Previously, it was shown that MDMD, in contrast to SDMD, is the MDD phenotype that associates with immune activation (Maes, Rachayon et al. 2022, Almulla, Abbas Abo Algon et al. 2024).

Using mCRP as a solitary biomarker distinguished MDD patients from controls, though with modest accuracy (58%). Incorporating albumin and transferrin raised accuracy above 72%, while the addition of serum IL-17A produced additional gains. By contrast, pCRP had no discriminatory value (Almulla, Kitov et al. 2025, Maes, Niu et al. 2025). These findings show that mCRP has greater relevance than pCRP, but negative APPs remain the strongest biomarkers of the immune-inflammatory response in MDD or SDMD.

The current study highlights that mCRP is most effective as part of a biomarker panel for MDD when combined with other immune-inflammatory markers such as serum albumin, transferrin and IL-17A. Likewise, a large part of the variance in OSOD was explained by the regression on mCRP (not pCRP), the negative APPs and IL-17A. Past work demonstrated a significant accuracy for negative APPs in melancholia, with sensitivity of 72% and specificity of 92% (Maes, Vandewoude et al. 1991, Maes 1995). These data align with recent findings confirming serum IL-17A as a biomarker of MDD (Maes and Carvalho 2018, Moulton, Malys et al. 2024) although other studies reported limited or sex-specific effects (Saraykar, Cao et al. 2018, Xie, Lai et al. 2023, Tsuboi, Sakakibara et al. 2024)

In contrast, Wessa et al. claimed that pCRP ≥ 3.0 mg/L defines “inflammatory depression” in about 30% of patients (Wessa, Janssens et al. 2024). Our data refute this stance, showing that pCRP is not even increased in MDD and, as explained below, is a metabolic biomarker. Even more important, employing mCRP coupled with negative APPs and IL-17A identifies around 63.7% of MDD patients with a reasonable accuracy. Thus, the conclusion that 30% of MDD patients show “inflammatory MDD” is inadequate because around 63% of those patients show an immune profile reminiscent of a smoldering immune response. In this respect, a recent study showed that 78.8% of MDD patients exhibit at least one NIMETOX abnormality (Maes, Jirakran et al. 2025). Furthermore, if one considers that the other phenotype of MDD, namely SDMD, often exhibits suppression of CIRS activities, one may conclude that most MDD patients, if not all, show phenotype-specific immune alterations. Thus, stratifying patients using this inadequate pCRP criterion and using this as an indicant to treat MDD with anti-inflammatory medications (Wessa et al., 2024) is fundamentally inaccurate.

### Determinants of CRP isoforms

The second major finding of this study indicates that mCRP is predicted by pCRP, BMI, the M1 macrophage profile and ACEs. In contrast, pCRP was predicted by BMI, ACEs, and M1 macrophages (positive) and CIRS (negative) profiles. As reviewed in the introduction, pCRP dissociates in serum into mCRP during increased inflammatory and oxidative load. This may also explain the difference between both CRP isoforms in their association with different immune profiles. Thus, while pCRP was associated with the M1 macrophage profile, which is known to induce the positive AP response in the liver, it was also inversely associated with the CIRS profile, which is known to inhibit the AP response (Maes et al., 2025). Other predictors of both pCRP and mCRP encompassed metabolic indices, including BMI, waist circumference and MetS ranking. Nevertheless, the relationship between pCRP and metabolic variables were more pronounced than those with mCRP. The findings support the notion that pCRP in the lower concentration ranges (1-10 mg/L,(Almulla, Kitov et al. 2025) is indicative of metabolic dysregulation rather than immune-inflammation processes associated with depression (Almulla, Kitov et al. 2025, Maes, Niu et al. 2025). Importantly, this study found that the association between mCRP and depression (diagnosis or severity) was independent from metabolic variables and pCRP. These findings indicate that mCRP, in contrast to pCRP, is a disease-specific biomarker of MDD and MDMD.

Both serum pCRP and mCRP were significantly predicted by ACEs. The latter also impacted the negative APP response (Maes, Niu et al. 2025), reflecting their broader impact on NIMETOX pathways (Maes, Almulla et al. 2025). As a consequence, ACEs play a critical role in the AP response established in MDD or MDMD. Prior studies showed that ACEs elevate CRP and MetS risk later in life (Danese, Pariante et al. 2007, Danese, Moffitt et al. 2009, Lin, Neylan et al. 2016, Iob, Lacey et al. 2020, Balaji and Sankaranarayanan 2023, Zagaria, Fiori et al. 2024, O’Shields, Slavich et al. 2025). Furthermore, the accuracy for MDD improved when ACEs were included along with mCRP and both negative APPs.

### mCRP as an inflammatory biomarker

mCRP enhances immune-inflammatory processes via various mechanisms. For example, mCRP promotes platelet aggregation, vascular activation, enhanced expression of adhesion molecules on endothelial cells, neutrophil migration, NK-cell activity, and activation of the complement system (Molins, Peña et al. 2011, Zeinolabediny, Kumar et al. 2021, Lazarut-Nistor and Slevin 2025). These actions promote thrombosis and vascular inflammation, phenomena also observed in MDD (Tonhajzerova, Sekaninova et al. 2020). Concurrently, mCRP enhances the expression of IL-6, IL-8, TNF-α, VCAM-1, COX-2, and MMP-13 via NF-κB signaling and promotes the generation of reactive oxygen species (Ruiz-Fernández, Gonzalez-Rodríguez et al. 2021, Olson, Hornick et al. 2023). Moreover, mCRP interacts with key immune pathways thereby activating microglia and inflammasomes via iNOS, COX-2, and NLRP3, thereby amplifying neuroinflammation in brain tissues (Bartra, Vuraić et al. 2025). The observed effects align closely with the immune-inflammatory, M1 macrophage and Th-17 patterns identified in MDMD (Maes, Rachayon et al. 2022, Almulla, Abbas Abo Algon et al. 2024), offering substantial evidence that mCRP plays a role in the immune signature associated with MDD and MDMD (Hornick and Potempa 2023).

In adaptive immunity, mCRP interacts with cholesterol in CD4+ T cell membranes, modifying TCR conformation to a primed state that increases IFN-γ release (Zhou, Chen et al. 2023). It indirectly activates T cells via monocytes by upregulating CD80, which subsequently engages CD80/CD28 co-stimulation. This pathway is inhibited by Belatacept, thereby affirming its specificity (Thomé, Limmer et al. 2025). Furthermore, neurodegenerative and autoimmune models highlight the pathogenic potential of mCRP. In Alzheimer’s disease, mCRP inhibits ApoE expression and disrupts ApoE–LRP1 binding, consequently heightening neuronal vulnerability (Na, Yang et al. 2023). In stroke models, mCRP is identified in endothelial cells and neurons within peri-infarct regions, associated with toxic inflammatory and angiogenic responses (Slevin, Matou-Nasri et al. 2010, Lazarut-Nistor and Slevin 2025). In lupus nephritis, immune complexes formed between mCRP and anti-mCRP antibodies intensify inflammation, interacting with anti-dsDNA and anti-C1q antibodies (Sjöwall, Bengtsson et al. 2003, Trouw, Groeneveld et al. 2004, Jönsen, Gunnarsson et al. 2007, Jakuszko, Krajewska et al. 2017). The findings suggest that mCRP may have detrimental effects as a neurodegenerative and inflammatory driver.

Interestingly, diminished albumin resulting from the adverse acute phase response may cause elevated mCRP levels. Albumin serves as the primary plasma transporter for lysophosphatidylcholine (LPC), which has cytotoxic properties and activates the immune response. Consequently, reduced albumin levels may increase free LPC bioavailability, facilitating the dissociation of pCRP into mCRP (Durço, Conceição et al. 2023).

## Limitations

This research presents several limitations. The cross-sectional design limits the ability to draw causal conclusions regarding the relationships among elevated mCRP, immune activation, negative APPs, and the severity of depression. Longitudinal investigations are required to determine whether mCRP changes precede, coincide with, or follow the clinical course of MDD. Second, the study population consisted exclusively of Chinese patients with MDD, which may limit the generalizability of the findings to other ethnic or cultural groups. Replication in diverse populations is therefore essential. Third, although mCRP was examined in combination with immune-inflammatory markers, future research should also evaluate its interaction with additional NIMETOX components, particularly oxidative and nitrosative stress pathways, and atherogenicity profiles (Maes, Almulla et al. 2025).

## Conclusion

This work provides the first evidence that serum mCRP is significantly elevated in MDD, most notably in MDMD, independent of BMI, medication status, and pCRP. The association of mCRP with M1 activation, Th-17 signaling, and reduced negative APPs defines a distinct immune-inflammatory phenotype in MDD largely overlapping with MDMD. These findings demonstrate that mCRP is more relevant than pCRP for characterizing MDD/MDMD and support its integration into precision psychiatry approaches.

## Data Availability

The corresponding author (MM) will provide access to the dataset supporting this study upon reasonable request, subsequent to a comprehensive data review.

## Acknowledgements

Not Applicable

## Ethical approval and consent to participate

The Research Ethics Committee of Sichuan Provincial People’s Hospital in Chengdu, China, granted approval for the study (Ethics Approval No. 2024-203). All participants submitted written informed consent before enrollment.

## Author’s contributions

All authors made equal contributions to the research and endorsed the final manuscript.

## References

1. Al-Marwani, S., A. Batieha, Y. Khader, M. El-Khateeb, H. Jaddou and K. Ajlouni (2023). “Association between albumin and depression: a population-based study.” BMC Psychiatry 23(1): 780.

2. Alberti, K. G., R. H. Eckel, S. M. Grundy, P. Z. Zimmet, J. I. Cleeman, K. A. Donato, J. C. Fruchart, W. P. James, C. M. Loria and S. C. Smith, Jr. (2009). “Harmonizing the metabolic syndrome: a joint interim statement of the International Diabetes Federation Task Force on Epidemiology and Prevention; National Heart, Lung, and Blood Institute; American Heart Association; World Heart Federation; International Atherosclerosis Society; and International Association for the Study of Obesity.” Circulation 120(16): 1640–1645.

3. Almulla, A. F., A. Abbas Abo Algon, C. Tunvirachaisakul, H. K. Al-Hakeim and M. Maes (2024). “T helper-1 activation via interleukin-16 is a key phenomenon in the acute phase of severe, first-episode major depressive disorder and suicidal behaviors.” J Adv Res 64: 171–181.

4. Almulla, A. F., A. A. A. Algon and M. Maes (2024). “Adverse childhood experiences and recent negative events are associated with activated immune and growth factor pathways, the phenome of first episode major depression and suicidal behaviors.” Psychiatry Res 334: 115812.

5. Almulla, A. F., S. Kitov, T. Deneva, M.-F. Kitova, L. Kitova, K. Stoyanova, D. Stoyanov and M. Maes (2025). “C-reactive protein is not a biomarker of depression severity in drug-naïve obese patients with metabolic syndrome.” medRxiv: 2025.2007.2008.25331082.

6. Almulla, A. F., Y. Thipakorn, A. Vasupanrajit, A. A. Abo Algon, C. Tunvirachaisakul, A. A. Hashim Aljanabi, G. Oxenkrug, H. K. Al-Hakeim and M. Maes (2022). “The tryptophan catabolite or kynurenine pathway in major depressive and bipolar disorder: A systematic review and meta-analysis.” Brain, Behavior, & Immunity - Health 26: 100537.

7. Ambrus, L. and S. Westling (2019). “Inverse association between serum albumin and depressive symptoms among drug-free individuals with a recent suicide attempt.” Nord J Psychiatry 73(4-5): 229–232.

8. Balaji, S. and A. Sankaranarayanan (2023). “The association between adverse childhood experiences and metabolic syndrome in severe mental illness: A literature review.” Australas Psychiatry 31(3): 381–388.

9. Bartra, C., K. Vuraić, Y. Yuan, S. Codony, H. Valdés-Quiroz, C. Casal, M. Slevin, L. Máquez-Kisinousky, A. M. Planas, C. Griñán-Ferré, M. Pallàs, C. Morisseau, B. D. Hammock, S. Vázquez, C. Suñol and C. Sanfeliu (2025). “Microglial pro-inflammatory mechanisms induced by monomeric C-reactive protein are counteracted by soluble epoxide hydrolase inhibitors.” Int Immunopharmacol 155: 114644.

10. Bernstein, D. P., J. A. Stein, M. D. Newcomb, E. Walker, D. Pogge, T. Ahluvalia, J. Stokes, L. Handelsman, M. Medrano, D. Desmond and W. Zule (2003). “Development and validation of a brief screening version of the Childhood Trauma Questionnaire.” Child Abuse Negl 27(2): 169–190.

11. Caprio, V., L. Badimon, M. Di Napoli, W. H. Fang, G. R. Ferris, B. Guo, R. S. Iemma, D. Liu, Y. Zeinolabediny and M. Slevin (2018). “pCRP-mCRP Dissociation Mechanisms as Potential Targets for the Development of Small-Molecule Anti-Inflammatory Chemotherapeutics.” Front Immunol 9: 1089.

12. Danese, A., T. E. Moffitt, H. Harrington, B. J. Milne, G. Polanczyk, C. M. Pariante, R. Poulton and A. Caspi (2009). “Adverse childhood experiences and adult risk factors for age-related disease: depression, inflammation, and clustering of metabolic risk markers.” Arch Pediatr Adolesc Med 163(12): 1135–1143.

13. Danese, A., C. M. Pariante, A. Caspi, A. Taylor and R. Poulton (2007). “Childhood maltreatment predicts adult inflammation in a life-course study.” Proc Natl Acad Sci U S A 104(4): 1319–1324.

14. Douglas, K. M., A. J. Taylor and P. G. O’Malley (2004). “Relationship Between Depression and C-Reactive Protein in a Screening Population.” Biopsychosocial Science and Medicine 66(5): 679–683.

15. Durço, A. O., L. S. R. Conceição and D. S. Souza (2023). “Albumin: to be, or not to be, a buffer, that is the question.” Journal of Applied Physiology 135(1): 201–201.

16. Gao, N., P. Yuan, Z. M. Tang, J. G. Lei, Z. R. Yang, M. Ahmed, Z. Y. Yao, D. Liang, Y. Wu and H. Y. Li (2024). “Monomeric C-reactive protein is associated with severity and prognosis of decompensated hepatitis B cirrhosis.” Front Immunol 15: 1407768.

17. Gregg, L. P., T. Carmody, D. Le, G. Martins, M. Trivedi and S. S. Hedayati (2020). “A Systematic Review and Meta-Analysis of Depression and Protein-Energy Wasting in Kidney Disease.” Kidney Int Rep 5(3): 318–330.

18. Hamilton, M. (1959). “The assessment of anxiety states by rating.” British journal of medical psychology.

19. Hamilton, M. (1960). “A rating scale for depression.” Journal of neurology, neurosurgery, and psychiatry 23(1): 56.

20. Hornick, M. G. and L. A. Potempa (2023). “Monomeric C-reactive protein as a biomarker for major depressive disorder.” Front Psychiatry 14: 1325220.

21. Iob, E., R. Lacey and A. Steptoe (2020). “The long-term association of adverse childhood experiences with C-reactive protein and hair cortisol: Cumulative risk versus dimensions of adversity.” Brain Behav Immun 87: 318–328.

22. Jakuszko, K., M. Krajewska, K. Kościelska-Kasprzak, M. Myszka, A. Sebastian, K. Gniewek, P. Wiland and M. Klinger (2017). “Antibodies against monomeric C-reactive protein - a promising biomarker of lupus nephritis?” Clin Biochem 50(13-14): 756–762.

23. Jha, M. K., M. Leboyer, C. M. Pariante and A. H. Miller (2025). “Should Inflammation Be a Specifier for Major Depression in the DSM-6?” JAMA Psychiatry 82(6): 549–550.

24. Jönsen, A., I. Gunnarsson, B. Gullstrand, E. Svenungsson, A. Bengtsson, O. Nived, I. Lundberg, L. Truedsson and G. Sturfelt (2007). “Association between SLE nephritis and polymorphic variants of the CRP and FcγRIIIa genes.” Rheumatology 46(9): 1417–1421.

25. Khan, A., D. Leonard, L. Defina, C. E. Barlow, B. Willis and E. S. Brown (2020). “Association between C Reactive Protein and Depression in a Population of Healthy Adults: The Cooper Center Longitudinal Study.” Journal of Investigative Medicine 68(5): 1019–1023.

26. Lazarut-Nistor, A. and M. Slevin (2025) “Beyond the Biomarker: Monomeric CRP as a Driver of Multisystem Pathology in Rheumatoid Arthritis.” International Journal of Molecular Sciences 26 DOI: 10.3390/ijms26178227.

27. Lin, J. E., T. C. Neylan, E. Epel and A. O’Donovan (2016). “Associations of childhood adversity and adulthood trauma with C-reactive protein: A cross-sectional population-based study.” Brain Behav Immun 53: 105–112.

28. Maes, M. (1993). “A review on the acute phase response in major depression.” Rev Neurosci 4(4): 407–416.

29. Maes, M. (1995). “Evidence for an immune response in major depression: a review and hypothesis.” Prog Neuropsychopharmacol Biol Psychiatry 19(1): 11–38.

30. Maes, M., A. F. Almulla, Z. You and Y. Zhang (2025). “Neuroimmune, metabolic and oxidative stress pathways in major depressive disorder.” Nat Rev Neurol.

31. Maes, M. and A. F. Carvalho (2018). “The Compensatory Immune-Regulatory Reflex System (CIRS) in Depression and Bipolar Disorder.” Mol Neurobiol 55(12): 8885–8903.

32. Maes, M., K. Jirakran, L. d. O. Semeão, A. P. Michelin, A. K. Matsumoto, F. F. Brinholi, D. S. Barbosa, C. Tivirachaisakul, A. F. Almulla, D. Stoyanov and Y. Zhang (2025). “Key factors underpinning neuroimmune-metabolic-oxidative (NIMETOX) major depression in outpatients: paraoxonase 1 activity, reverse cholesterol transport, increased atherogenicity, protein oxidation, and differently expressed cytokine networks.” medRxiv: 2025.2003.2002.25323183.

33. Maes, M., M. Niu, X. Zhang, J. Li, D. Stoyanov, B. Zhou, A. F. Almulla and Y. Zhang (2025). “The negative acute phase response and not serum C-reactive protein is a major biomarker of major depression: a precision nomothetic psychiatry study.” medRxiv: 2025.2008.2015.25333787.

34. Maes, M., M. Rachayon, K. Jirakran, P. Sodsai, S. Klinchanhom, P. Gałecki, A. Sughondhabirom and A. Basta-Kaim (2022) “The Immune Profile of Major Dysmood Disorder: Proof of Concept and Mechanism Using the Precision Nomothetic Psychiatry Approach.” Cells 11 DOI: 10.3390/cells11071183.

35. Maes, M., S. Scharpe, E. Bosmans, M. Vandewoude, E. Suy, W. Uyttenbroeck, W. Cooreman, C. Vandervorst and J. Raus (1992). “Disturbances in acute phase plasma proteins during melancholia: additional evidence for the presence of an inflammatory process during that illness.” Prog Neuropsychopharmacol Biol Psychiatry 16(4): 501–515.

36. Maes, M., M. Vandewoude, S. Scharpé, L. De Clercq, W. Stevens, L. Lepoutre and C. Schotte (1991). “Anthropometric and biochemical assessment of the nutritional state in depression: evidence for lower visceral protein plasma levels in depression.” J Affect Disord 23(1): 25–33.

37. Maes, M., A. Vasupanrajit, K. Jirakran, B. Zhou, C. Tunvirachaisakul and A. F. Almulla (2023). “Major depression is not an inflammatory disorder: depletion of the compensatory immunoregulatory system is a hallmark of a mild depression phenotype.” medRxiv: 2023.2012.2014.23299942.

38. Maes, M., A. Vasupanrajit, K. Jirakran, B. Zhou, C. Tunvirachaisakul and A. F. Almulla (2025). “Simple dysmood disorder, a mild subtype of major depression, is not an inflammatory condition: Depletion of the compensatory immunoregulatory system.” Journal of Affective Disorders 375: 75–85.

39. Molins, B., E. Peña, R. De La Torre and L. Badimón (2011). “Monomeric C-reactive protein is prothrombotic and dissociates from circulating pentameric C-reactive protein on adhered activated platelets under flow.” Cardiovascular research 92 **2**: 328–337.

40. Moraes, J. B., M. Maes, D. S. Barbosa, T. Z. Ferrari, M. K. S. Uehara, A. F. Carvalho and S. O. V. Nunes (2017). “Elevated C-reactive Protein Levels in Women with Bipolar Disorder may be Explained by a History of Childhood Trauma, Especially Sexual Abuse, Body Mass Index and Age.” CNS Neurol Disord Drug Targets 16(4): 514–521.

41. Moulton, C. D., M. Malys, C. W. P. Hopkins, A. S. Rokakis, A. H. Young and N. Powell (2024). “Activation of the interleukin-23/Th17 axis in major depression: a systematic review and meta-analysis.” Eur Arch Psychiatry Clin Neurosci.

42. Na, H., J. B. Yang, Z. Zhang, Q. Gan, H. Tian, I. M. Rajab, L. A. Potempa, Q. Tao and W. Q. Qiu (2023). “Peripheral apolipoprotein E proteins and their binding to LRP1 antagonize Alzheimer’s disease pathogenesis in the brain during peripheral chronic inflammation.” Neurobiology of Aging 127: 54–69.

43. O’Shields, J. D., G. M. Slavich and O. Mowbray (2025). “Adverse childhood experiences, inflammation, and depression: evidence of sex-and stressor specific effects in a nationally representative longitudinal sample of U.S. adolescents.” Psychol Med 55: e140.

44. Olson, M. E., M. G. Hornick, A. Stefanski, H. R. Albanna, A. Gjoni, G. D. Hall, P. C. Hart, I. M. Rajab and L. A. Potempa (2023). “A biofunctional review of C-reactive protein (CRP) as a mediator of inflammatory and immune responses: differentiating pentameric and modified CRP isoform effects.” Front Immunol 14: 1264383.

45. Raison, C. L., R. E. Rutherford, B. J. Woolwine, C. Shuo, P. Schettler, D. F. Drake, E. Haroon and A. H. Miller (2013). “A randomized controlled trial of the tumor necrosis factor antagonist infliximab for treatment-resistant depression: the role of baseline inflammatory biomarkers.” JAMA Psychiatry 70(1): 31–41.

46. Ruiz-Fernández, C., M. Gonzalez-Rodríguez, V. Francisco, I. M. Rajab, R. Gómez, J. Conde, F. Lago, J. Pino, A. Mobasheri, M. A. Gonzalez-Gay, A. Mera, L. A. Potempa and O. Gualillo (2021). “Monomeric C reactive protein (mCRP) regulates inflammatory responses in human and mouse chondrocytes.” Lab Invest 101(12): 1550–1560.

47. Saraykar, S., B. Cao, L. S. Barroso, K. S. Pereira, L. Bertola, M. Nicolau, J. D. Ferreira, N. S. Dias, E. L. Vieira, A. L. Teixeira, A. P. M. Silva and B. S. Diniz (2018). “Plasma IL-17A levels in patients with late-life depression.” Braz J Psychiatry 40(2): 212–215.

48. Sheehan, D. V., Y. Lecrubier, K. H. Sheehan, P. Amorim, J. Janavs, E. Weiller, T. Hergueta, R. Baker and G. C. Dunbar (1998). “The Mini-International Neuropsychiatric Interview (M.I.N.I.): the development and validation of a structured diagnostic psychiatric interview for DSM-IV and ICD-10.” J Clin Psychiatry 59 **Suppl 20**: 22–33;quiz 34-57.

49. Sjöwall, C., A. A. Bengtsson, G. Sturfelt and T. Skogh (2003). “Serum levels of autoantibodies against monomeric C-reactive protein are correlated with disease activity in systemic lupus erythematosus.” Arthritis Res Ther 6(2): R87.

50. Slevin, M., S. Matou-Nasri, M. Turu, A. Luque, N. Rovira, L. Badimon, S. Boluda, L. Potempa, C. Sanfeliu, N. De Vera and J. Krupinski (2010). “Modified C-Reactive Protein Is Expressed by Stroke Neovessels and Is a Potent Activator of Angiogenesis In Vitro.” Brain Pathology 20(1): 151–165.

51. Spielberger, C., R. Gorsuch, R. Lushene, P. Vagg and G. Jacobs (1983). “Manual for the State-Trait Anxiety Inventory; Palo Alto, CA, Ed.” Palo Alto: Spielberger.

52. Thiele, J. R., J. Habersberger, D. Braig, Y. Schmidt, K. Goerendt, V. Maurer, H. Bannasch, A. Scheichl, K. J. Woollard, E. von Dobschütz, F. Kolodgie, R. Virmani, G. B. Stark, K. Peter and S. U. Eisenhardt (2014). “Dissociation of pentameric to monomeric C-reactive protein localizes and aggravates inflammation: in vivo proof of a powerful proinflammatory mechanism and a new anti-inflammatory strategy.” Circulation 130(1): 35–50.

53. Thomé, J., J. Limmer, T. Z. Brose, J. Zeller, N. Chevalier, A. L. Schäfer, L. Schneider, M. Lind, T. Christmann, M. Dreck, S. Kreuzaler, D. Braig, K. Peter, F. Pankratz and S. U. Eisenhardt (2025). “C-reactive protein induced T cell activation is an indirect monocyte-dependent mechanism involving the CD80/CD28 pathway.” Front Immunol 16: 1622865.

54. Tonhajzerova, I., N. Sekaninova, L. Bona Olexova and Z. Visnovcova (2020) “Novel Insight into Neuroimmune Regulatory Mechanisms and Biomarkers Linking Major Depression and Vascular Diseases: The Dilemma Continues.” International Journal of Molecular Sciences 21 DOI: 10.3390/ijms21072317.

55. Trouw, L. A., T. W. Groeneveld, M. A. Seelen, J. M. Duijs, I. M. Bajema, F. A. Prins, U. Kishore, D. J. Salant, J. S. Verbeek and C. Van Kooten (2004). “Anti-C1q autoantibodies deposit in glomeruli but are only pathogenic in combination with glomerular C1q-containing immune complexes.” The Journal of clinical investigation 114(5): 679–688.

56. Tsuboi, H., H. Sakakibara, Y. Minamida-Urata, H. Tsujiguchi, A. Hara, K. Suzuki, S. Miyagi, M. Nakamura, C. Takazawa, T. Kannon, J. Zhao, Y. Shimizu, A. Shibata, A. Ogawa, F. Suzuki, Y. Kambayashi, T. Konoshita, A. Tajima and H. Nakamura (2024). “Serum TNFα and IL-17A levels may predict increased depressive symptoms: findings from the Shika Study cohort project in Japan.” BioPsychoSocial Medicine 18(1): 20.

57. Vasupanrajit, A., M. Maes, K. Jirakran and C. Tunvirachaisakul (2024). “Complex Intersections Between Adverse Childhood Experiences and Negative Life Events Impact the Phenome of Major Depression.” Psychol Res Behav Manag 17: 2161–2178.

58. Wessa, C., J. Janssens, V. Coppens, K. El Abdellati, E. Vergaelen, S. van den Ameele, C. Baeken, D. Zeeuws, Y. Milaneschi, F. Lamers, B. Penninx, S. Claes, M. Morrens and L. De Picker (2024). “Efficacy of inflammation-based stratification for add-on celecoxib or minocycline in major depressive disorder: Protocol of the INSTA-MD double-blind placebo-controlled randomised clinical trial.” Brain Behav Immun Health 41: 100871.

59. Xie, X.-h., W.-t. Lai, S.-x. Xu, M. Di Forti, J.-y. Zhang, M.-m. Chen, L.-h. Yao, P. Wang, K.-k. Hao and H. Rong (2023). “Hyper-inflammation of astrocytes in patients of major depressive disorder: Evidence from serum astrocyte-derived extracellular vesicles.” Brain, Behavior, and Immunity 109: 51–62.

60. Zagaria, A., V. Fiori, M. Vacca, C. Lombardo, C. M. Pariante and A. Ballesio (2024). “Inflammation as a mediator between adverse childhood experiences and adult depression: A meta-analytic structural equation model.” J Affect Disord 357: 85–96.

61. Zeinolabediny, Y., S. Kumar and M. Slevin (2021). “Monomeric C-Reactive Protein - A Feature of Inflammatory Disease Associated With Cardiovascular Pathophysiological Complications?” In Vivo 35(2): 693–697.

62. Zhao, X., Zhang, Yalin, Li, Longfei, Zhou, Yunfei, Li, Hezhan, & Yang, Shichang (2005). “Reliability and validity of the Chinese version of the Childhood Abuse Questionnaire.” Chinese Clinical Rehabilitation Journal(20): 105–107.

63. Zhou, L., S. J. Chen, Y. Chang, S. H. Liu, Y. F. Zhou, X. P. Huang, Y. X. Hua, H. An, S. H. Zhang, I. Melnikov, Z. A. Gabbasov, Y. Wu and S. R. Ji (2023). “Monomeric C-reactive protein evokes TCR Signaling-dependent bystander activation of CD4+ T cells.” Mol Immunol 157: 158–166.

